# A Spatially Validated Machine-Learning Framework for Mapping Alpha-Gal Syndrome Exposure Potential in Illinois

**DOI:** 10.64898/2026.07.24.26358812

**Authors:** Abrar Hussain, Nohra Mateus-Pinilla, Rebecca L. Smith

**Affiliations:** Department of Pathobiology, College of Veterinary Medicine, University of Illinois Urbana-Champaign, Urbana, Illinois, USA; Illinois Natural History Survey, Prairie Research Institute, University of Illinois Urbana-Champaign, Champaign, Illinois, USA; Department of Biomedical and Translational Sciences, Carle Illinois College of Medicine, University of Illinois Urbana-Champaign, Illinois, USA

**Keywords:** *Amblyomma americanum*, alpha-gal syndrome, ehrlichiosis, machine learning, spatial cross-validation, exposure-risk mapping, Illinois

## Abstract

The lone star tick, *Amblyomma americanum*, is closely associated with alpha-gal syndrome (AGS), an emerging tick-associated allergic condition. However, heterogeneous tick surveillance and limited clinical AGS data complicate the assessment of geographic exposure. We investigated whether ecological modeling and epidemiological surveillance could identify areas of potentially elevated AGS exposure across Illinois. Using data from 102 Illinois counties, eight statistical and machine-learning models were evaluated through geographic cross-validation to predict the relative abundance of lone star ticks. Random forest and XGBoost predictions were separately combined with reported ehrlichiosis rates to construct county-level AGS exposure-risk indices. Wildland-urban interface (WUI) population was examined in sensitivity analyses. Random forest achieved near-minimum prediction error while XGBoost demonstrated the strongest rank correlation. Both indices showed substantial concordance with CDC suspected-AGS categories across 97 counties (Spearman ρ = 0.710 and 0.721). At the highest category threshold, odds ratios per 0.10-unit index increase were 5.02 and 3.63, respectively. By integrating ecological predictions with epidemiological surveillance, we establish a reproducible, spatially explicit framework for prioritizing AGS surveillance, targeting interventions, and guiding future epidemiological investigations across Illinois counties.

## 1. Introduction

Alpha-gal syndrome (AGS) is an emerging tick bite-associated allergic condition characterized by delayed reactions to mammalian meat and other products containing galactose-α-1,3-galactose (alpha-gal) [1,2]. In the United States, the lone star tick (*Amblyomma americanum*) is the primary tick species associated with alpha-gal sensitization. The Centers for Disease Control and Prevention (CDC) estimated that as many as 450,000 people in the United States may be affected by AGS, although its actual burden remains uncertain [3]. National surveillance of suspected AGS has identified geographic concentrations in the southern, Midwestern, and Mid-Atlantic United States, including Illinois [4]. Identifying areas of potentially elevated exposure is therefore important for improving surveillance, clinical awareness, and public health prevention.

Despite growing recognition of AGS, detailed geographic information on clinically confirmed cases and tick-exposure locations remains limited. The CDC’s published county-level distribution is based on positive alpha-gal-specific antibody testing rather than confirmed clinical diagnoses, leaving uncertainty about the geographic burden of AGS [4]. An Illinois survey found that only 32% of physicians knew the cause of AGS, and fewer than 18% were aware of available diagnostic testing [5]. Among surveyed Illinois farmers, 42% reported knowledge of lone star ticks [6], while a subsequent comparison of Illinois stakeholder surveys identified broader gaps in tick and tick-borne disease knowledge and prevention practices [7]. In Illinois, the 2025 Tracking Infectious Cases Knowledgeably (TICK) Act emphasized the need for AGS awareness and education for healthcare providers [8]. Meanwhile, differences in tick collection methods and surveillance coverage complicate geographic comparisons of tick relative abundance, highlighting the need for integrated ecological and epidemiological approaches to assess AGS exposure.

Statistical and machine-learning approaches have increasingly been applied to predict tick distributions and habitat suitability by integrating climatic, vegetation, land-use, and host-related variables [9–11]. Such approaches can estimate ecological patterns across counties where direct surveillance observations are incomplete or inconsistent. However, spatial dependence among neighboring counties may inflate predictive performance when geographic structure is ignored. Geographic cross-validation and spatial modeling provide complementary approaches for evaluating regional predictions and accounting for spatial dependence [12].

Ecological predictions of lone star tick relative abundance alone cannot fully describe opportunities for human exposure. Human ehrlichiosis provides additional epidemiologic information because *A. americanum* transmits ehrlichiosis-causing pathogens, and the CDC has documented broad geographic overlap between ehrlichiosis and suspected AGS [4,13]. The wildland-urban interface (WUI), where residential development meets or intermingles with natural vegetation, may also indicate environments where human-tick encounters are possible [14]. Combining these indicators with ecological predictions could provide a more informative estimate of the relative representation of potential AGS exposure, provided their distinct meanings and uncertainties are recognized. Transparent weighting and sensitivity analyses are particularly important when combining indicators that measure related but distinct processes [15].

In this study, we used statistical and machine-learning models, including spatial extensions, to predict geographic variation in the relative abundance of lone star ticks across Illinois based on environmental and host-habitat characteristics. We evaluated model performance across 102 counties using geographically separated validation groups and integrated predictions from selected high-performing models with epidemiological evidence of human-tick exposure to develop county-level AGS exposure-risk scores. We assessed the geographic concordance between these scores and CDC-suspected AGS categories to determine whether integrating ecological and epidemiological evidence could identify geographic patterns of elevated AGS exposure risk in areas where direct disease surveillance remains limited.

## 2. Materials and Methods

The overall analytical workflow for developing and evaluating county-level AGS exposure-risk indices is illustrated in Figure 1.

**Figure 1.**
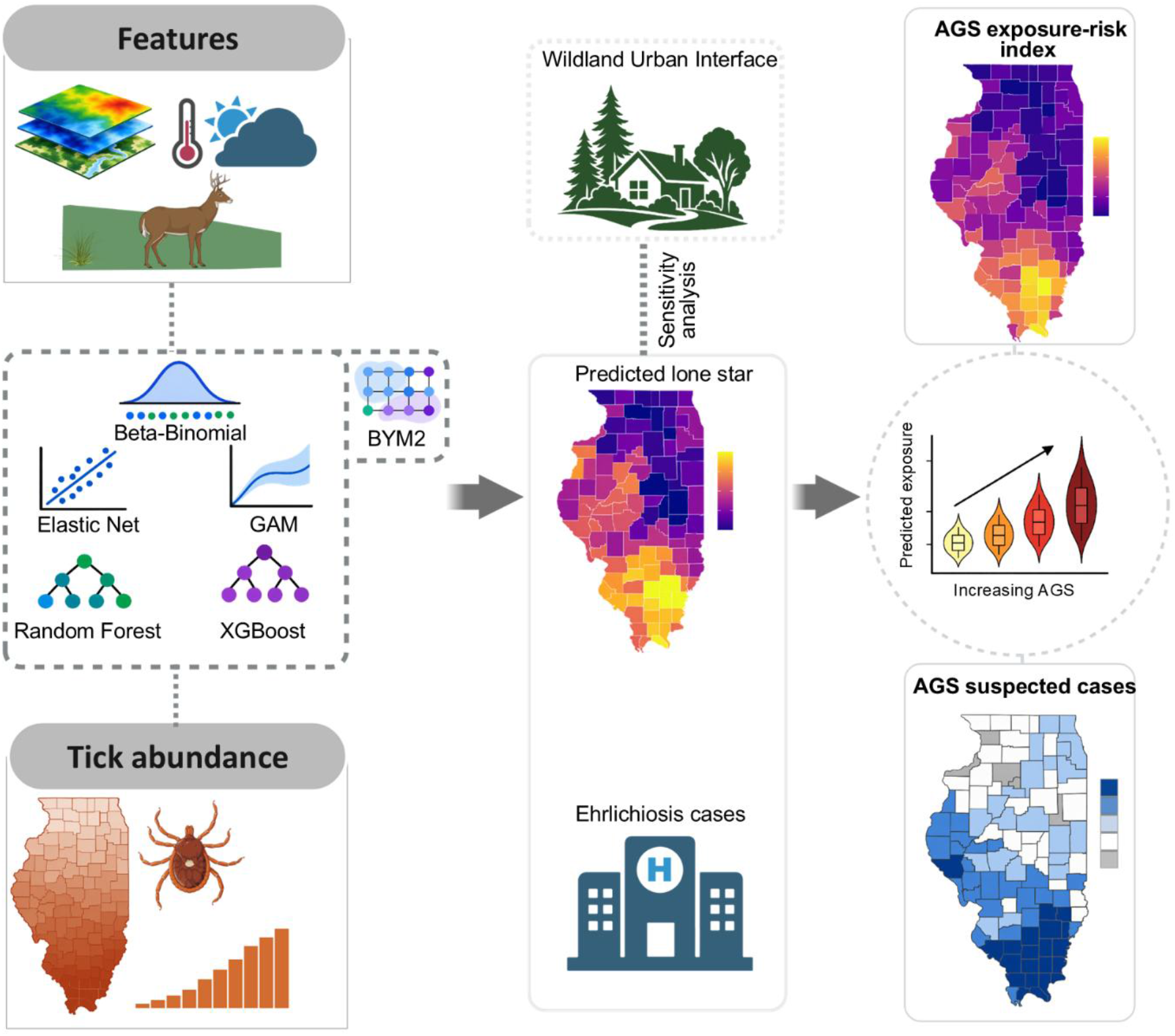
Overview of the workflow for developing and evaluating alpha-gal syndrome (AGS) exposure-risk indices in Illinois. Ecological predictors and tick surveillance informed five baseline models, of which the three best-performing were also compared with their BYM2 spatial-extension versions. Selected predictions were combined with ehrlichiosis incidence and, in sensitivity analyses, wildland-urban interface (WUI) population. Indices were evaluated against CDC suspected-AGS categories.

### 2.1 Tick surveillance and ecological covariates

The analysis included all 102 counties in Illinois. County-level tick surveillance data were obtained from the Illinois Department of Public Health (IDPH) for 2022-2024 [16]. The proportion of collected ticks identified as *A. americanum* was calculated by dividing the number of lone star ticks by the total number of collected ticks across all species. This proportion was used as a measure of relative abundance within surveillance collections, rather than relying on raw tick counts, which may vary with surveillance intensity. This proportion served as the primary outcome for statistical and machine-learning models. All counties had positive total tick counts, including 33 counties with no recorded lone star ticks.

We initially considered nine county-level ecological predictors selected to represent climatic conditions influencing tick survival and activity, vegetation and landscape characteristics associated with suitable tick habitat, and habitat suitability for important wildlife hosts. Five bioclimatic variables were derived from WorldClim historical monthly climate data based on the Climatic Research Unit Time Series (CRU TS) for 2022-2024 [17,18]: mean diurnal temperature range (BIO2), mean temperature of the wettest quarter (BIO8), mean temperature of the driest quarter (BIO9), precipitation seasonality (BIO15), and precipitation of the warmest quarter (BIO18). These variables capture temperature and moisture conditions relevant to tick development, survival, and seasonal activity. Land-cover predictors comprised forest and wetland coverage derived from the Annual National Land Cover Database (NLCD) for 2022-2024 [19,20], representing vegetation and moisture-related habitat conditions. White-tailed deer habitat suitability [21] was included because deer are important hosts for adult *A. americanum*, while forest-edge density was considered to capture habitat transitions that may influence tick and host distributions [22,23]. All predictors were summarized at the county level, and multicollinearity among the nine candidate predictors was assessed using pairwise Pearson correlation coefficients and variance inflation factors (VIFs) (Figure S1). Forest-edge density was excluded because it overlapped with other forest-related predictors, leaving eight ecological variables for model development (Supplementary material).

### 2.2. Predictive modeling and spatial validation

We used a two-stage modeling approach to evaluate county-level lone star tick relative abundance and determine whether incorporating explicit spatial dependence improved geographic predictions. First, five baseline statistical and machine-learning models were evaluated: beta-binomial regression, elastic-net binomial regression, generalized additive models (GAMs), random forest (RF), and extreme gradient boosting (XGBoost). Beta-binomial regression was implemented using the glmmTMB package to model lone star tick counts relative to other collected ticks while accommodating overdispersion [24]. Elastic-net binomial regression was implemented using glmnet, combining L1 (Lasso) and L2 (Ridge) regularization to account for correlated ecological predictors [25]. Generalized additive models (GAMs) were fitted using mgcv with a quasibinomial logit link and cubic regression splines to capture nonlinear ecological relationships [26]. Random forest regression was implemented using ranger, while extreme gradient boosting (XGBoost) used a squared-error regression objective to model complex nonlinear associations and interactions among predictors [27,28]. County total tick counts were incorporated through binomial count responses in beta-binomial and elastic-net models, observation weights in GAM and XGBoost, and sampling weights in RF.

Geographic predictive performance was evaluated using five-fold spatial cross-validation. Counties were assigned to five geographic groups using k-means clustering of projected county coordinates (EPSG:5070), independently of observed tick proportions (Table S1; Figure S2). In each iteration, one geographic group was withheld for evaluation, while the remaining four groups were used for model fitting and hyperparameter selection. Beta-binomial regression used training-fold-standardized ecological predictors. Elastic-net tuning evaluated five regularization mixing parameters, with the optimal penalty selected through inner geographic cross-validation. GAMs used cubic regression splines (k = 4), with smoothing parameters estimated by restricted maximum likelihood (REML). RF tuning evaluated 12 combinations of candidate predictors per split (mtry = 2, 4, 6, 8) and minimum terminal-node sizes (3, 5, 10). XGBoost tuning considered maximum tree depths of 1-3, learning rates of 0.03 and 0.10, and minimum child weights of 1 and 5, with early stopping to control model complexity. RF and XGBoost hyperparameters were selected using inner geographically held-out RMSE.

In the second stage, the three best-performing baseline models were extended using Besag-York-Mollié 2 (BYM2) spatial effects to assess whether accounting for spatial dependence improved geographic predictive performance [29]. County adjacency was defined using queen contiguity, in which counties sharing a boundary or vertex were considered neighbors. The spatial extensions incorporated spatially structured and unstructured county effects. For the machine-learning extensions, cross-fitted, logit-transformed predictions were incorporated into spatial regression models alongside BYM2 effects. Withheld county outcomes were excluded from spatial model fitting.

Together, the five baseline models and three spatial extensions yielded eight models for comparison. The same outer geographic partitions were used for all eight models, and predictions from the five withheld groups were combined to generate one out-of-fold (OOF) prediction per county and model. Predictive performance was assessed using root mean squared error (RMSE), mean absolute error (MAE), coefficient of determination (R^2^), and Spearman rank correlation between observed and OOF-predicted lone star tick proportions. Fold-specific performance was also examined to assess geographic variation in prediction accuracy and whether spatial extensions provided additional predictive benefit. For ecological interpretation, selected high-performing models were subsequently fitted using all 102 counties and examined through variable importance, accumulated local effects (ALE), and SHapley Additive exPlanations (SHAP) [30]. Full-data models were used exclusively for ecological interpretation, while predictive accuracy was evaluated using geographically held-out predictions. Additional implementation details are provided in the Supplementary material.

### 2.3. Development of AGS exposure-risk indices

County-level AGS exposure-risk indices were constructed by combining predicted relative abundance of lone star ticks with human ehrlichiosis surveillance data for 2022-2024 [31]. Ehrlichiosis was used as an epidemiological proxy for human-tick exposure because it shares an important tick vector with AGS and exhibits overlapping geographic distributions [4,13]. For each county ***i***, the annualized ehrlichiosis incidence per 100,000 residents was calculated as:

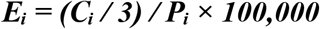

where ***C***_***i***_ represents the total reported ehrlichiosis cases during 2022-2024, and ***P***_***i***_ denotes the county population. To reduce the influence of highly skewed incidence rates, ***E***_***i***_ was transformed as log (1 + ***E***_***i***_) and then normalized to a 0-1 scale across all 102 counties via min-max scaling, yielding ***H***_***i***_.

For ecological prediction model ***m***, the primary AGS exposure-risk index was calculated as:

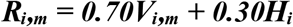

where ***V***_***i,m***_ represents the geographically out-of-fold predicted lone star tick proportion and ***H***_***i***_ represents the normalized ehrlichiosis incidence. The 70:30 weighting was predefined based on four considerations. First, AGS is a tick bite-associated allergic condition rather than a pathogen-mediated infection, making lone star tick ecology the primary biological indicator of potential exposure. Second, reported human ehrlichiosis cases provide only indirect evidence of human-tick encounters with *Ehrlichia*-positive lone star ticks, as they represent diagnosed and reported infections rather than the full extent of tick exposure. Third, ehrlichiosis cases are assigned by county of residence, which may differ from the actual location of tick exposure, limiting their geographic precision as an exposure proxy. Finally, the weights were specified independently of CDC-suspected AGS categories, which were reserved for external evaluation, thereby avoiding optimization against the same data used to assess geographic concordance. Accordingly, the 70% ecological and 30% epidemiological contributions reflect the biological relevance and limitations of the available indicators rather than an empirically established optimal ratio [4,13]. Weighting sensitivity was assessed using alternative ecological-to-epidemiological ratios of 50:50, 60:40, and 80:20 to evaluate the stability of county rankings relative to the primary 70:30 index. Additive aggregation allowed both components to contribute without requiring simultaneous high levels, consistent with composite-indicator methodology [15]. Index construction was performed independently for model-specific ecological predictions to preserve their geographic variation.

To evaluate the potential contribution of human contact opportunities, a sensitivity index was constructed to capture the proportion of each county’s population residing within the wildland-urban interface (WUI), derived from the Spatial Analysis For Conservation and Sustainability (SILVIS) version 4 dataset [14]. The alternative index was calculated as:

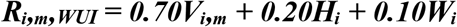

where ***W***_***i***_ represents the county population proportion residing in WUI areas, ranging from 0 to 1. The ecological component was maintained at 70%, while 10 percentage points were reassigned from the epidemiological component to the WUI indicator to examine the sensitivity of geographic rankings to an additional measure of potential human-tick contact. The WUI-adjusted index was calculated separately for each ecological model selected based on geographic cross-validation performance. Spearman rank correlations among the primary, alternative-weighted, and WUI-adjusted indices were calculated across all 102 counties to assess the stability of rankings. Detailed data processing and index implementation are provided in Supplementary material.

### 2.4. External evaluation using CDC suspected-AGS data

To assess geographic concordance between the developed AGS exposure-risk indices and observed suspected-AGS patterns, county-level categories were reconstructed from the published Centers for Disease Control and Prevention (CDC) map based on alpha-gal-specific immunoglobulin E (IgE) laboratory testing during 2017-2022 [4]. The map classified annual suspected-AGS rates per million residents into four ordered categories: zero, <11, 11-87, and >87, with a separate designation for counties where no testing was performed. The digitized categories were matched to Illinois counties using standardized county names. Five counties without testing information were excluded, leaving 97 counties for evaluation. These categories reflect suspected AGS identified through laboratory testing rather than the incidence of clinically confirmed disease. The CDC and surveillance datasets overlapped only in 2022; therefore, the analysis assessed geographic concordance rather than temporal agreement.

Geographic concordance between each AGS exposure-risk index and the ordered CDC categories was evaluated using Spearman rank correlation, with 95% confidence intervals estimated from 5,000 nonparametric bootstrap resamples of counties. Associations were further examined using cumulative logistic regression, with index values scaled to express odds ratios (ORs) per 0.10-unit increase. The proportional-odds assumption was assessed using likelihood-ratio tests of nominal effects. Because the assumption was not satisfied for the primary indices, non-proportional cumulative logit models were fitted, allowing associations to vary across the three cumulative category thresholds. Threshold-specific ORs with Wald 95% confidence intervals were estimated, and predicted probabilities of each CDC category were calculated across the observed index ranges. The same regression specification was applied to the wildland-urban interface (WUI) sensitivity indices for comparability.

### 2.5. Spatial sensitivity analysis

To examine whether incorporating neighboring-county information influenced geographic concordance with CDC suspected-AGS patterns, an exploratory spatial averaging analysis was performed for the primary and WUI sensitivity indices. Using queen-contiguous county adjacency, one-step spatial averaging assigned equal weight to each county’s original index (50%) and the mean index of its adjacent counties (50%). This approach incorporated local geographic context while retaining the original county-specific contribution. Spatial averaging was applied independently to each index using the original scores. Spearman rank correlations with CDC suspected-AGS categories were recalculated for the 97 counties with available testing information. Correlations before and after spatial averaging were compared, and changes in correlation coefficients (**Δρ**) were calculated to evaluate the sensitivity of geographic concordance to neighboring-county information.

## 3. Results

### 3.1. Lone star tick distribution and ecological patterns

Across 102 Illinois counties, surveillance records included 5,366 *Amblyomma americanum* ticks among 12,938 collected ticks. The mean county-level lone star tick proportion was 0.394 (SD = 0.373), with a median of 0.324 and a range of 0-1. Overall, 33 counties (32.4%) had no recorded lone star ticks. Geographic variation was pronounced, with lower proportions concentrated in northern Illinois and greater lone star tick relative abundance across much of central and southern Illinois (Figure 2). Mean observed proportions across the five geographic folds ranged from 0.089 to 0.660, reflecting substantial regional heterogeneity (Table S1). Ecological covariates exhibited distinct geographic patterns, with higher temperatures during the driest quarter (BIO9) and generally greater deer habitat suitability in southern Illinois, whereas forest and wetland coverage showed more localized spatial variation (Figure S3).

**Figure 2.**
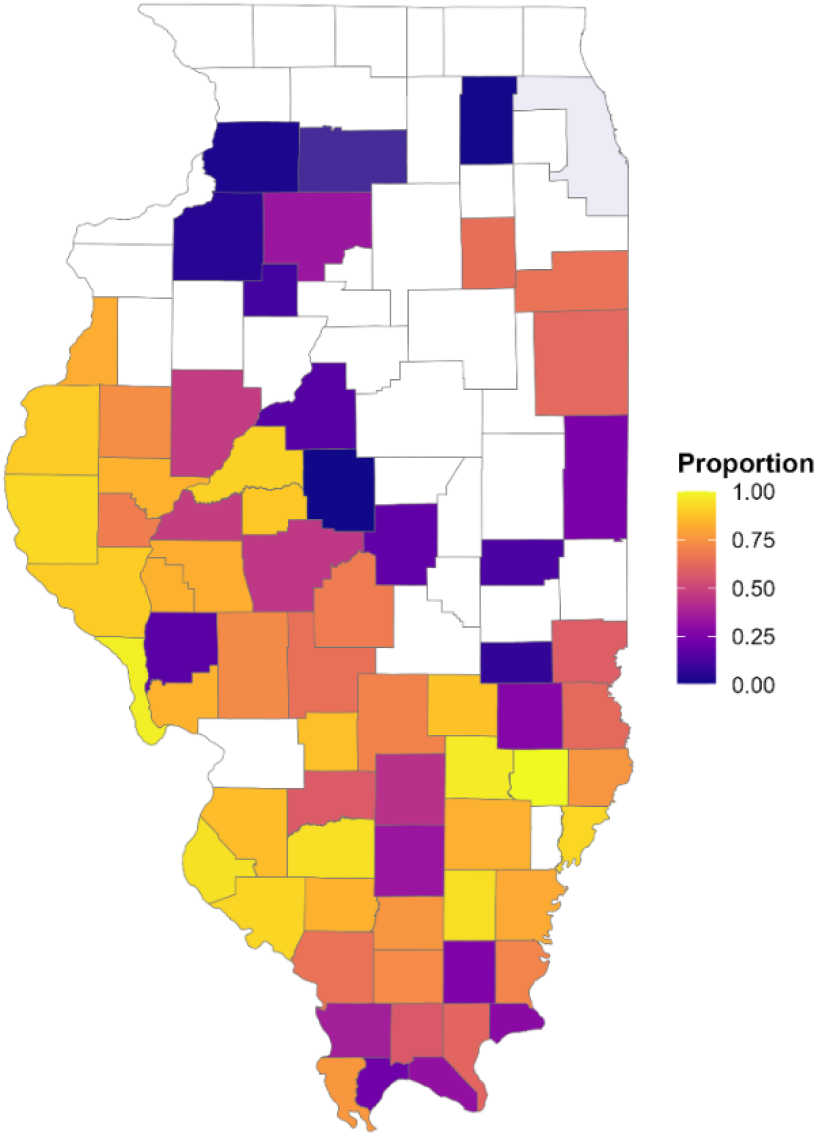
Geographic distribution of the observed proportion of collected ticks identified as *Amblyomma americanum* across 102 Illinois counties during 2022-2024. White counties indicate a zero proportion of lone star ticks in surveillance collections, rather than an absence of tick sampling.

### 3.2. Spatial predictive performance

Spatial cross-validation revealed differences in predictive accuracy and geographic generalizability among the eight models (Table S2). Random forest achieved near-minimum prediction error (RMSE = 0.316), comparable to XGBoost-BYM2 (0.313), while XGBoost demonstrated the strongest county-level rank correlation (ρ = 0.556) and lowest worst-fold RMSE. Although incorporating BYM2 spatial effects improved overall RMSE for some models, these extensions did not consistently improve performance across geographically withheld folds (Table S3). Comparisons of observed and predicted lone star tick proportion rank groups revealed some classification discrepancies for random forest and XGBoost (Figure S4).

Given the marginal improvement in overall RMSE from spatial extensions and their inconsistent benefits across geographic folds, random forest and XGBoost were selected for ecological interpretation and development of AGS exposure-risk indices based on their complementary predictive strengths and direct relationships with ecological covariates.

### 3.3. Ecological predictors and model interpretation

Both random forest (RF) and XGBoost consistently identified five leading ecological predictors: temperature of the driest quarter (BIO9), mean diurnal temperature range (BIO2), temperature of the wettest quarter (BIO8), deer habitat suitability, and forest cover (Figure S5). Temperature-related variables dominated both models, with BIO9 ranking highest in RF and BIO2 in XGBoost. Although their relative importance varied, these five predictors consistently contributed most to the model’s predictions.

Accumulated local effects (ALE) revealed broadly similar nonlinear ecological relationships across both models (Figure 3). Predicted lone star tick proportions increased with higher mean diurnal temperature ranges and warmer temperatures during the driest quarter. Deer habitat suitability showed a generally positive relationship, whereas the temperature of the wettest quarter exhibited more pronounced increases at the highest values. SHapley Additive exPlanations (SHAP) further supported these patterns, with higher temperature values and greater deer suitability generally contributing positively to county-level predictions, whereas lower values contributed negatively (Figure 3). Although the magnitude and shape of individual predictor effects varied between models, their overall ecological patterns were largely consistent.

**Figure 3.**
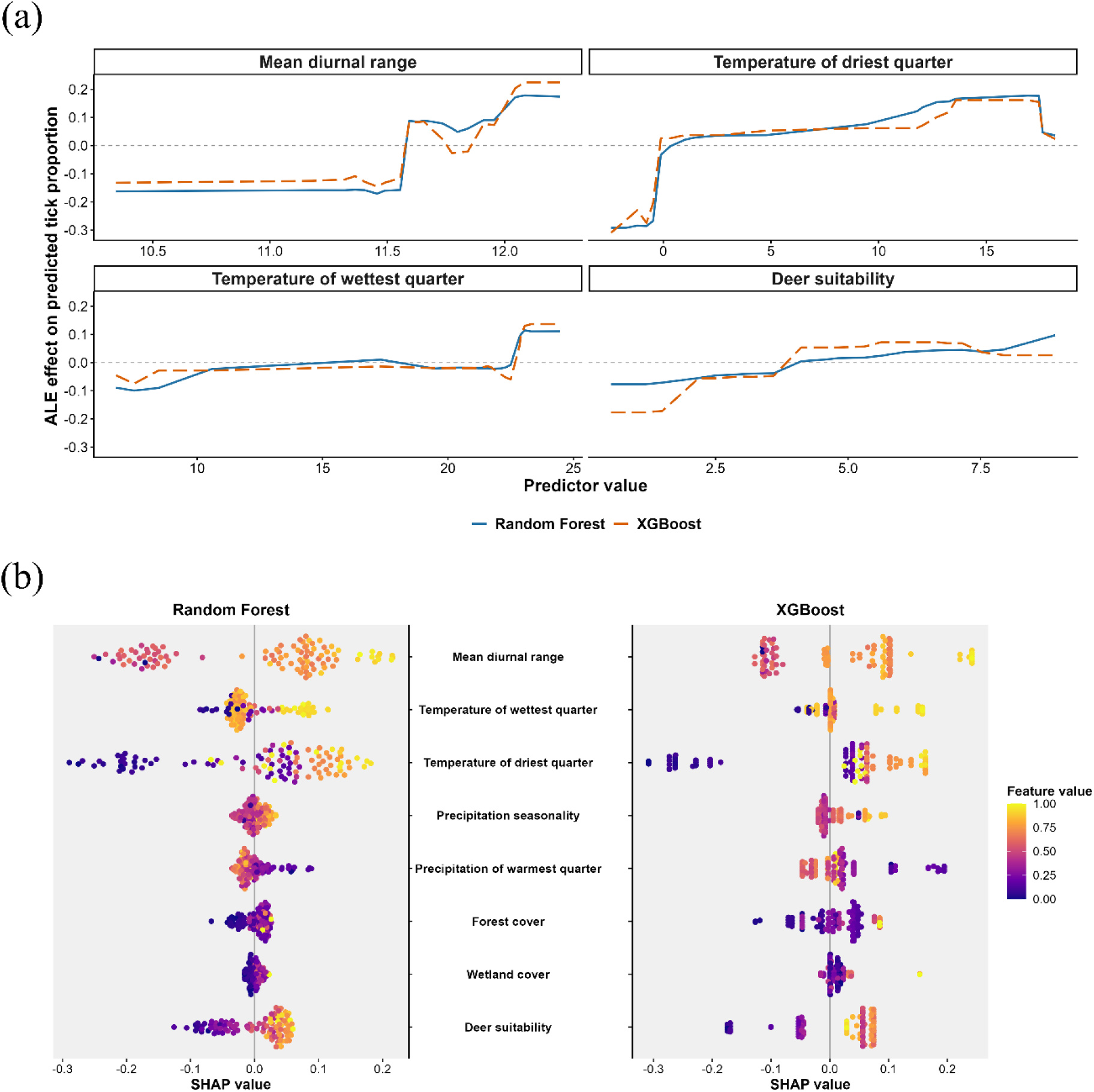
Ecological interpretation of random forest and XGBoost models. (a) Accumulated local effects of four ecological predictors (random forest: solid blue; XGBoost: dashed orange). (b) SHAP beeswarm plots showing county-level predictor contributions, colored by feature values.

### 3.4. Geographic distribution of AGS exposure-risk indices

The primary AGS exposure-risk indices, combining geographically cross-validated lone star tick predictions from RF or XGBoost (70%) with scaled ehrlichiosis incidence (30%), showed broadly consistent geographic patterns (Figure 4). Higher index scores were concentrated in southern Illinois, with Hamilton, Massac, Pope, Jefferson, and Franklin counties ranking among the five highest in both models. County rankings were strongly correlated between the RF and XGBoost indices (Spearman ρ = 0.952), although XGBoost produced a wider range of index values than RF.

**Figure 4.**
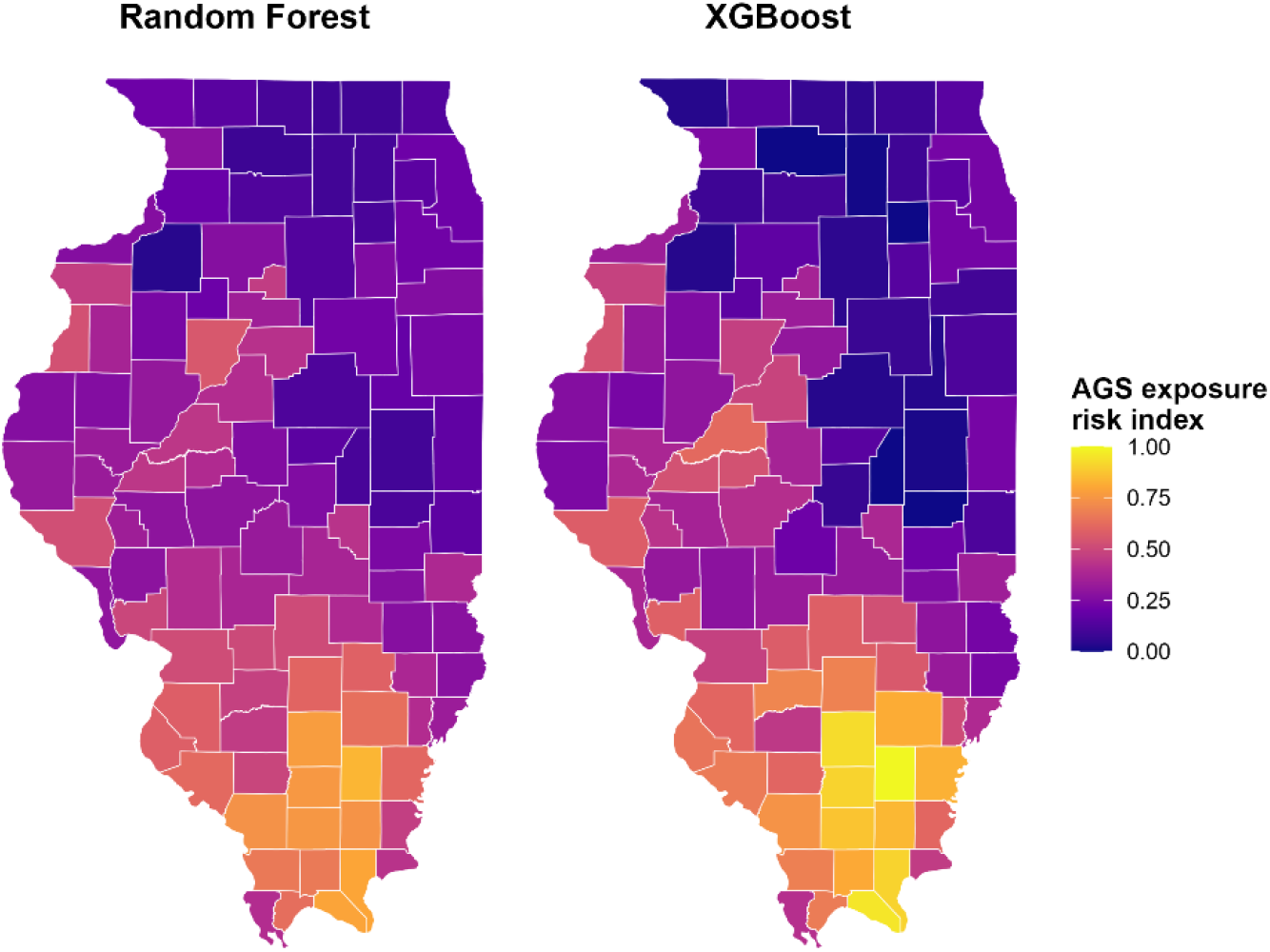
Geographic distribution of primary alpha-gal syndrome exposure-risk indices across Illinois. County-level indices derived from random forest and XGBoost predictions, combined with scaled ehrlichiosis incidence at a 70:30 weighting.

Incorporating wildland-urban interface (WUI) population in the 70:20:10 sensitivity indices preserved similar geographic patterns (Figure S6). Compared with their respective primary indices, county rankings remained highly correlated for RF (ρ = 0.995) and XGBoost (ρ = 0.997), with the same ten highest-ranked counties retained. Alternative ecological-to-ehrlichiosis weightings of 50:50, 60:40, and 80:20 also showed strong agreement with the primary 70:30 formulation (ρ = 0.984-0.996 for RF and 0.990-0.997 for XGBoost; Figure S7), indicating limited sensitivity of county prioritization to index weighting.

### 3.5 *Geographic* concordance *with CDC suspected-AGS categories*

Among 97 Illinois counties with available CDC suspected-AGS categories (Figure S8), both primary exposure-risk indices showed strong positive geographic concordance (Spearman ρ = 0.710 for random forest and 0.721 for XGBoost; both p < 0.001). Index scores generally increased across successive CDC categories (Figure 5). The proportional-odds assumption was not supported for either index, and threshold-specific ordinal regression showed increasing odds with higher index scores (Table S4). Predicted probabilities from the non-proportional ordinal models illustrated the corresponding shifts in CDC category membership as random forest and XGBoost index values increased (Figure 6). The strongest associations occurred at the highest category threshold, with odds ratios of 5.02 for random forest and 3.63 for XGBoost per 0.10-unit increase in the respective index.

**Figure 5.**
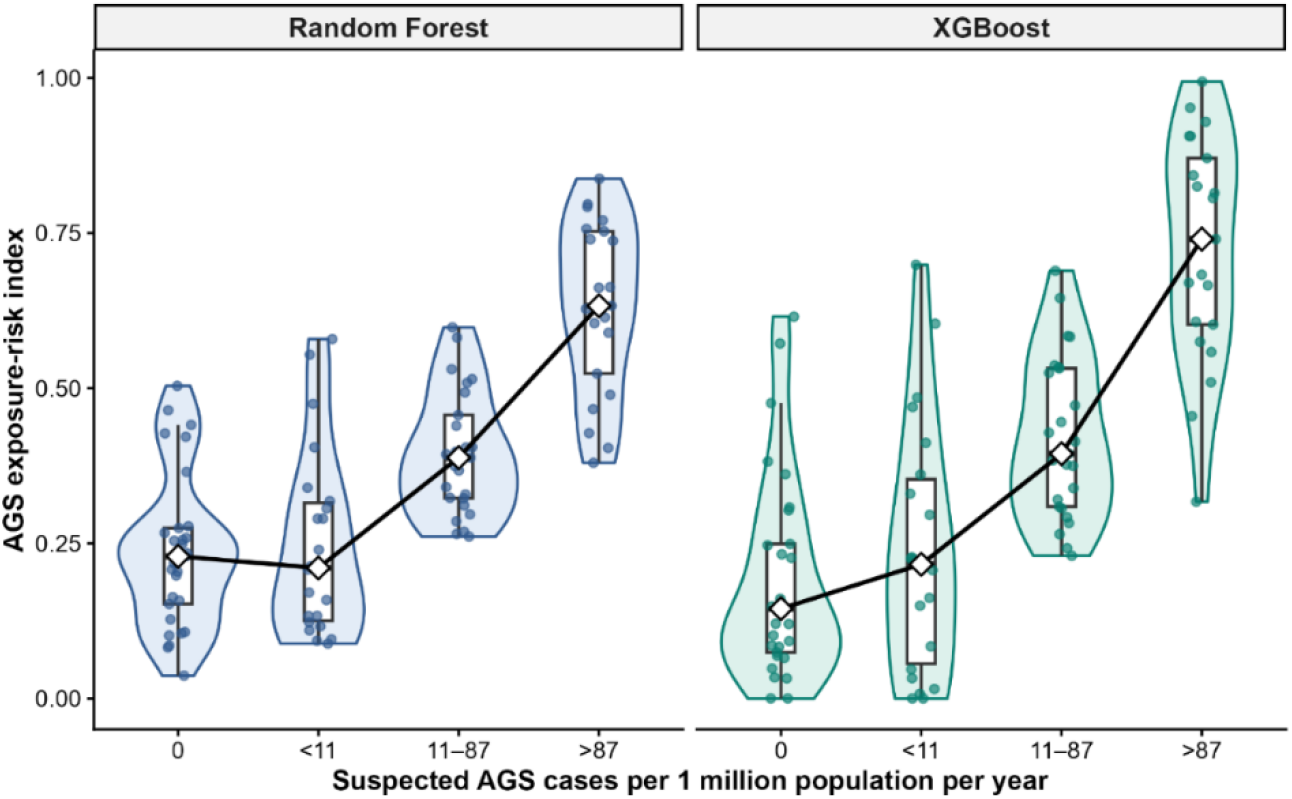
Distribution of primary AGS exposure-risk indices across CDC suspected-AGS categories. Violin and box plots show RF and XGBoost indices across four CDC categories (n = 97). Points represent counties, and diamonds indicate category means.

**Figure 6.**
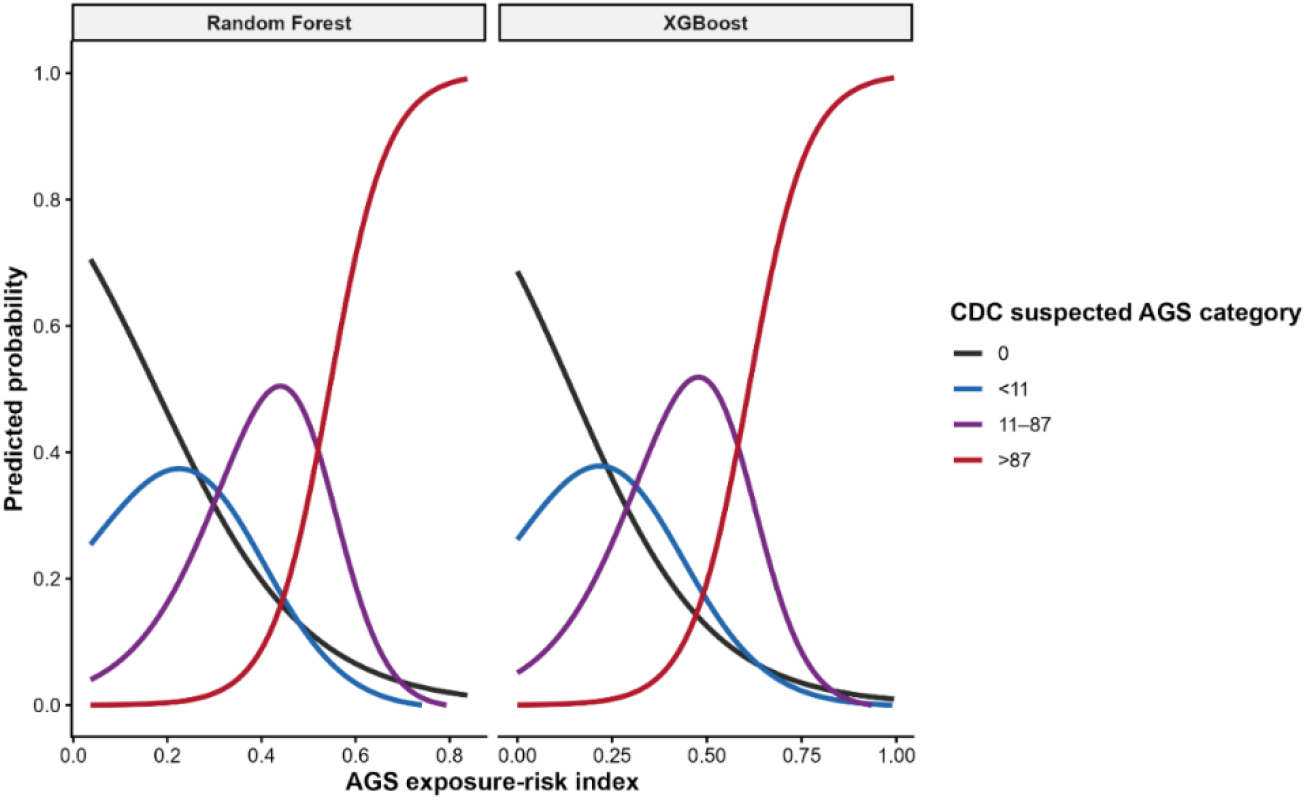
Predicted probabilities of CDC suspected-AGS categories across primary exposure-risk indices. Nonproportional ordinal regression curves illustrate category-specific probabilities across random forest and XGBoost index values.

Results remained consistent across sensitivity analyses. Incorporating wildland-urban interface (WUI) population yielded comparable correlations with CDC categories (ρ = 0.709 for random forest and 0.725 for XGBoost). Spatial averaging further increased correlations to 0.757 for random forest and 0.773 for XGBoost, indicating improved geographic concordance after incorporating neighboring county scores (Figure S9).

## 4. Discussion

A central challenge in understanding the geography of AGS is that ecological conditions favorable to its principal implicated tick species do not directly reveal where human sensitization or clinical disease occurs. This study addresses that gap by integrating spatially validated ecological predictions with an epidemiological indicator of human-tick encounters to construct county-level AGS exposure-risk indices for Illinois. The convergence of two machine-learning approaches on similar geographic priorities, together with their concordance with independently reported suspected-AGS patterns, provides evidence that heterogeneous surveillance data can be translated into a structured assessment of relative exposure potential. Previous national investigations established broad geographic overlap among suspected AGS, lone star ticks, and ehrlichiosis [4]. Our contribution extends this observation by developing quantitative county rankings based on geographically validated ecological predictions and epidemiological surveillance, which are evaluated against independent suspected AGS data.

The ecological coherence of the two models strengthens the biological interpretation of the resulting geographic patterns. Mean temperatures of the driest and wettest quarters, mean diurnal temperature range, deer habitat suitability, and forest cover consistently emerged as influential predictors, suggesting that lone star tick relative abundance reflects interconnected climatic and host-habitat conditions. Previous distribution models have emphasized the importance of climatic suitability, including moisture-related constraints, and have documented the complex environmental influences on the geographic range of *A. americanum* [32,33]. The nonlinear responses observed in both models indicate that these ecological relationships vary across environmental gradients. The importance of deer habitat suitability is biologically plausible, as white-tailed deer are major hosts for adult lone star ticks, and deer-targeted acaricide treatment reduced host-seeking ticks by more than 90% [34].

The modeling results also illustrate why geographic prediction requires more rigorous evaluation than conventional model-fit assessments. Models can reproduce existing spatial gradients while performing poorly when transferred to counties with different environmental characteristics or surveillance compositions. Spatially structured cross-validation addresses this concern by evaluating predictions in withheld geographic regions, an approach recommended for environmentally dependent datasets [12]. Although the spatially extended XGBoost model achieved the lowest overall prediction error, this advantage did not translate into consistently better performance across geographic folds. Random forest and XGBoost therefore offered complementary strengths in prediction accuracy and county-level ranking, while retaining direct relationships between ecological predictors and model outputs. These findings demonstrate why model selection for spatial risk assessment should consider geographic transferability alongside overall predictive accuracy.

Combining predicted lone star tick relative abundance with ehrlichiosis surveillance links an ecological indicator to an indirect measure of human-tick encounters, while recognizing that AGS and ehrlichiosis have distinct disease mechanisms. Their geographic overlap, previously documented in national surveillance, provides a biological rationale for integrating these information streams without assuming that their underlying disease processes are equivalent [4]. County rankings remained stable across alternative weighting scenarios and after incorporating the proportion of residents living within the WUI. The stability of county rankings following WUI integration further demonstrates the robustness of the proposed framework to alternative estimates of the relative representation of potential human-tick contact.

The correspondence with CDC suspected AGS categories provides an additional, human-health-oriented assessment of the framework. Higher exposure-risk indices were associated with higher suspected-AGS categories, and threshold-specific analyses indicated particularly strong differentiation at the highest category boundary. However, this agreement requires careful interpretation. The CDC categories were derived from laboratory testing rather than clinically confirmed AGS, and alpha-gal-specific immunoglobulin E (IgE) positivity does not necessarily indicate symptomatic disease [1,4,35]. But the strong concordance with independently derived CDC suspected-AGS categories provides epidemiological support for the geographic patterns identified by the framework, while improved concordance following spatial averaging further highlights the importance of regional ecological and epidemiological processes.

Several methodological choices strengthened the reliability of our findings. Geographic cross-validation assessed model transferability; comparisons across eight models and alternative index formulations examined sensitivity to analytical choices; and CDC-suspected AGS categories were reserved for external evaluation rather than weight optimization. Nevertheless, the interpretation of these findings remains subject to limitations in the underlying surveillance data. Relative abundance was estimated from the species composition of tick collections rather than standardized measures of tick density or human biting frequency, and geographic variation in collection practices may influence these estimates [36]. Seasonal variation in tick activity and finer-scale exposure information were not available in the aggregated surveillance data, while ehrlichiosis reporting and alpha-gal testing may be influenced by healthcare access and diagnostic practices. County-level aggregation and differences in surveillance periods also limit fine-scale and temporal interpretation. Future integration of standardized longitudinal tick surveillance, human-tick encounter data, and clinically confirmed AGS outcomes could further strengthen the framework and enable more comprehensive evaluation of its geographic predictive performance.

These findings are particularly relevant to the evolving public health response to AGS in Illinois. The state’s 2025 Tracking Infectious Cases Knowledgeably (TICK) Act established provisions for AGS education and awareness among communities, healthcare providers, and local health departments [8], while proposed legislation in 2026 sought to establish formal statewide and county-level case reporting [37]. The resulting county-level indices provide an evidence-based foundation for prioritizing targeted tick surveillance, guiding clinician education, and identifying geographic areas requiring further epidemiological investigation. Such applications are especially relevant given documented gaps in healthcare provider awareness and confidence in diagnosing AGS [38]. The framework could also support a staged surveillance strategy in which initial county rankings guide data collection, newly collected observations update ecological predictions, and emerging clinical information is used for independent evaluation. The framework may also be transferable to other regions where ecological surveillance is available, but direct AGS surveillance remains incomplete. Its principal value lies in providing a transparent and testable foundation on which more comprehensive surveillance and prevention efforts can be developed.

## 5. Conclusion

This study demonstrates that integrating spatially validated ecological predictions with epidemiological surveillance provides a reproducible framework for mapping AGS exposure potential across Illinois. The strong geographic concordance with CDC-suspected AGS categories, the consistency between the random forest and XGBoost indices, and the stability across sensitivity analyses support the framework’s utility for identifying geographic priorities. These findings provide an evidence-based foundation for targeted tick surveillance, public health awareness, and future epidemiological investigations, with potential applications in other regions where direct AGS surveillance remains limited.

## Data Availability

Analysis code is available at https://github.com/abrarhussain95/Alpha-Gal-Risk-in-Illinois. County-level tick surveillance and human ehrlichiosis data for 2022-2024 were obtained through web scraping of publicly accessible Illinois Department of Public Health dashboards. Environmental datasets are available from their respective sources. CDC suspected-AGS categories were derived from publicly available CDC data. Redistribution of the IDPH datasets remains subject to confirmation of applicable data-sharing permissions.

## Acknowledgments

We sincerely thank the researchers and surveillance personnel who contributed to documenting tick occurrence records in Illinois. We also gratefully acknowledge the Illinois Department of Public Health for making data available.

## CRediT authorship contribution statement

**Abrar Hussain:** Conceptualization, Methodology, Data Curation, Visualization, Formal analysis, Software, Writing-original draft, Writing-review & editing; **Nohra Mateus-Pinilla**: Conceptualization, Writing-review & editing; **Rebecca L. Smith**: Conceptualization, Writing-review & editing, Supervision.

## Data Availability Statement

Data and analysis code used in this study are publicly available at: https://github.com/abrarhussain95/Alpha-Gal-Risk-in-Illinois

