## Supplementary material for "A Spatially Validated Machine-Learning Framework for Mapping Alpha-Gal Syndrome Exposure Potential in Illinois"

This supplementary material accompanies the manuscript *"A Spatially Validated Machine-Learning Framework for Mapping Alpha-Gal Syndrome Exposure Potential in Illinois"* and includes additional methodological details, four supplementary tables (Tables S1-S4), and nine supplementary figures (Figures S1-S9).

#### Methods

##### Ecological covariates

Monthly WorldClim historical temperature and precipitation data based on CRU TS 4.09 were processed at 2.5 arc-minute spatial resolution for 2022-2024. Bioclimatic indices were derived from the monthly climate surfaces, summarized by county, and averaged across years. Annual NLCD land-cover data were used to calculate county-level forest and wetland coverage based on classes 41-43 and 90 and 95, respectively. The 2022 deer habitat-suitability raster was summarized by county. VIFs for the eight retained predictors were subsequently examined to assess remaining multicollinearity. Correlation and VIF diagnostics are presented in Supplementary Figure S1.

##### Model implementation and computational details

Geographic folds were generated using 100 random initializations, a maximum of 100 iterations, and a fixed random seed of 123. Elastic-net mixing parameters were evaluated at 0, 0.25, 0.50, 0.75, and 1.00, with the regularization penalty selected by minimizing inner cross-validated binomial deviance. Generalized additive models additionally incorporated shrinkage-based term selection. Random forest models used 500 trees during tuning, and 1,000 trees for outer-fold predictions, with county tick counts normalized by their training-set mean before being applied as sampling weights. XGBoost used observation and predictor subsampling fractions of 0.8, up to 500 boosting rounds, and early stopping after 30 rounds without validation improvement. The final number of boosting rounds was determined from the median optimal iteration across inner folds, with a minimum of 10 rounds.

The BYM2 spatial components were specified with scaled spatial effects and sum-to-zero constraints. Penalized-complexity priors were assigned to spatial precision (parameters 1 and 0.01) and the spatial mixing parameter (parameters 0.5 and 0.5), which controls the relative contribution of spatially structured and unstructured variation. Beta-binomial likelihoods used county-total tick counts as denominators. For the RF-BYM2 and XGBoost-BYM2 extensions, predictions for training counties were generated via inner geographic cross-fitting, whereas predictions for withheld counties were obtained from the corresponding outer-fold machine-learning models. Predicted proportions were restricted to the range 0.0001-0.9999 before the logit transformation to avoid undefined values at the boundaries. Withheld county outcomes were treated as missing during spatial fitting.

Additional diagnostics included calibration intercepts and slopes and residual spatial autocorrelation assessed using Moran's I with 999 Monte Carlo permutations. Variable importance scores were independently normalized to a maximum of 100. Accumulated local effects (ALE) were calculated using 20 intervals for BIO2, BIO8, BIO9, and deer habitat suitability. SHapley Additive exPlanations (SHAP) were estimated using tree-based or permutation-based approaches, as appropriate to each model, to quantify individual predictor contributions.

*Epidemiological data processing and index implementation*

County-level ecological predictions, ehrlichiosis surveillance records, and population data were matched by county, with common scaling limits applied across the model-specific indices. For the WUI component, census blocks classified as interface or intermix in the SILVIS 2020 WUI dataset (version 4) were identified. The population residing in these blocks was summed for each county and divided by the county's total census-block population. This population-weight approach represented the proportion of residents living in WUI areas rather than the proportion of county land classified as WUI.

#### Tables

**Table S1.** Distribution of lone star tick relative abundance across five geographic cross-validation folds in Illinois.

| Fold | Counties (n) | Mean tick proportion | Median tick proportion | Counties with zero lone star ticks (n) |
|---|---|---|---|---|
| 1 | 19 | 0.102 | 0.000 | 13 |
| 2 | 17 | 0.089 | 0.000 | 11 |
| 3 | 19 | 0.660 | 0.823 | 1 |
| 4 | 22 | 0.391 | 0.261 | 7 |
| 5 | 25 | 0.623 | 0.705 | 1 |

**Table S2.** Spatial cross-validation performance of eight models

| Model | RMSE | MAE | $R^2$ | Spearman ρ |
|---|---|---|---|---|
| XGBoost-BYM2 | 0.313 | 0.253 | 0.287 | 0.541 |
| Random forest | 0.316 | 0.266 | 0.275 | 0.532 |
| RF-BYM2 | 0.321 | 0.253 | 0.253 | 0.533 |
| XGBoost | 0.323 | 0.262 | 0.244 | 0.556 |
| Beta-binomial BYM2 | 0.34 | 0.261 | 0.162 | 0.481 |
| Beta-binomial | 0.354 | 0.27 | 0.09 | 0.481 |
| GAM | 0.374 | 0.288 | −0.018 | 0.43 |
| Elastic net | 0.442 | 0.395 | −0.420 | −0.042 |

**Table S3.** Fold-specific spatial cross-validation performance of eight models.

| Model | Geographic fold | RMSE | MAE | Spearman rho |
|---|---|---|---|---|
| XGBoost-BYM2 | 1 | 0.221 | 0.146 | 0.381 |
| XGBoost-BYM2 | 2 | 0.236 | 0.207 | 0.115 |
| XGBoost-BYM2 | 3 | 0.495 | 0.462 | -0.039 |
| XGBoost-BYM2 | 4 | 0.274 | 0.243 | 0.806 |

| | | | | |
|---|---|---|---|---|
| XGBoost-BYM2 | 5 | 0.271 | 0.217 | -0.006 |
| Random forest (RF) | 1 | 0.218 | 0.204 | 0.489 |
| Random forest (RF) | 2 | 0.419 | 0.357 | -0.052 |
| Random forest (RF) | 3 | 0.392 | 0.352 | 0.03 |
| Random forest (RF) | 4 | 0.267 | 0.23 | 0.706 |
| Random forest (RF) | 5 | 0.269 | 0.217 | 0.261 |
| RF-BYM2 | 1 | 0.223 | 0.139 | 0.427 |
| RF-BYM2 | 2 | 0.246 | 0.197 | -0.003 |
| RF-BYM2 | 3 | 0.527 | 0.484 | 0.007 |
| RF-BYM2 | 4 | 0.259 | 0.232 | 0.805 |
| RF-BYM2 | 5 | 0.266 | 0.221 | 0.213 |
| XGBoost | 1 | 0.24 | 0.177 | 0.3 |
| XGBoost | 2 | 0.378 | 0.306 | 0.029 |
| XGBoost | 3 | 0.357 | 0.319 | 0.067 |
| XGBoost | 4 | 0.274 | 0.227 | 0.643 |
| XGBoost | 5 | 0.348 | 0.283 | -0.019 |
| Beta-binomial BYM2 | 1 | 0.238 | 0.119 | 0.087 |
| Beta-binomial BYM2 | 2 | 0.238 | 0.204 | -0.143 |
| Beta-binomial BYM2 | 3 | 0.551 | 0.513 | -0.211 |
| Beta-binomial BYM2 | 4 | 0.267 | 0.237 | 0.749 |
| Beta-binomial BYM2 | 5 | 0.309 | 0.237 | -0.072 |
| Beta-binomial | 1 | 0.242 | 0.12 | 0.004 |
| Beta-binomial | 2 | 0.237 | 0.202 | -0.143 |
| Beta-binomial | 3 | 0.558 | 0.518 | -0.211 |
| Beta-binomial | 4 | 0.263 | 0.23 | 0.722 |
| Beta-binomial | 5 | 0.362 | 0.276 | -0.159 |
| GAM | 1 | 0.217 | 0.115 | 0.302 |
| GAM | 2 | 0.416 | 0.352 | -0.158 |
| GAM | 3 | 0.527 | 0.483 | -0.128 |

| | | | | |
|---|---|---|---|---|
| GAM | 4 | 0.297 | 0.229 | 0.642 |
| GAM | 5 | 0.36 | 0.28 | 0.158 |
| Elastic net | 1 | 0.448 | 0.431 | -0.242 |
| Elastic net | 2 | 0.513 | 0.502 | NA |
| Elastic net | 3 | 0.28 | 0.238 | 0.271 |
| Elastic net | 4 | 0.436 | 0.379 | -0.462 |
| Elastic net | 5 | 0.49 | 0.429 | 0.007 |

**Table S4**. Associations between AGS exposure-risk indices and CDC suspected-AGS categories across 97 Illinois counties.

| Model | CDC category threshold | Primary OR (95% CI) | WUI-adjusted OR (95% CI) |
|---|---|---|---|
| Random forest | 0 vs >0 | 1.87 (1.34-2.60) | 1.92 (1.37-2.69) |
| Random forest | <11 vs ≥11 | 2.59 (1.79-3.74) | 2.74 (1.88-4.01) |
| Random forest | ≤87 vs >87 | 5.02 (2.53-9.97) | 5.89 (2.80-12.42) |
| XGBoost | 0 vs >0 | 1.73 (1.33-2.24) | 1.75 (1.34-2.29) |
| XGBoost | <11 vs ≥11 | 2.14 (1.60-2.86) | 2.21 (1.64-2.97) |
| XGBoost | ≤87 vs >87 | 3.63 (2.05-6.43) | 3.95 (2.15-7.27) |

*OR = odds ratio per 0.10-unit increase in index score; CI = confidence interval. Primary weighting: 70:30. WUI-adjusted weighting: 70:20:10. All associations p < 0.001. Thresholds use suspected AGS cases per million population annually.*

### Figures

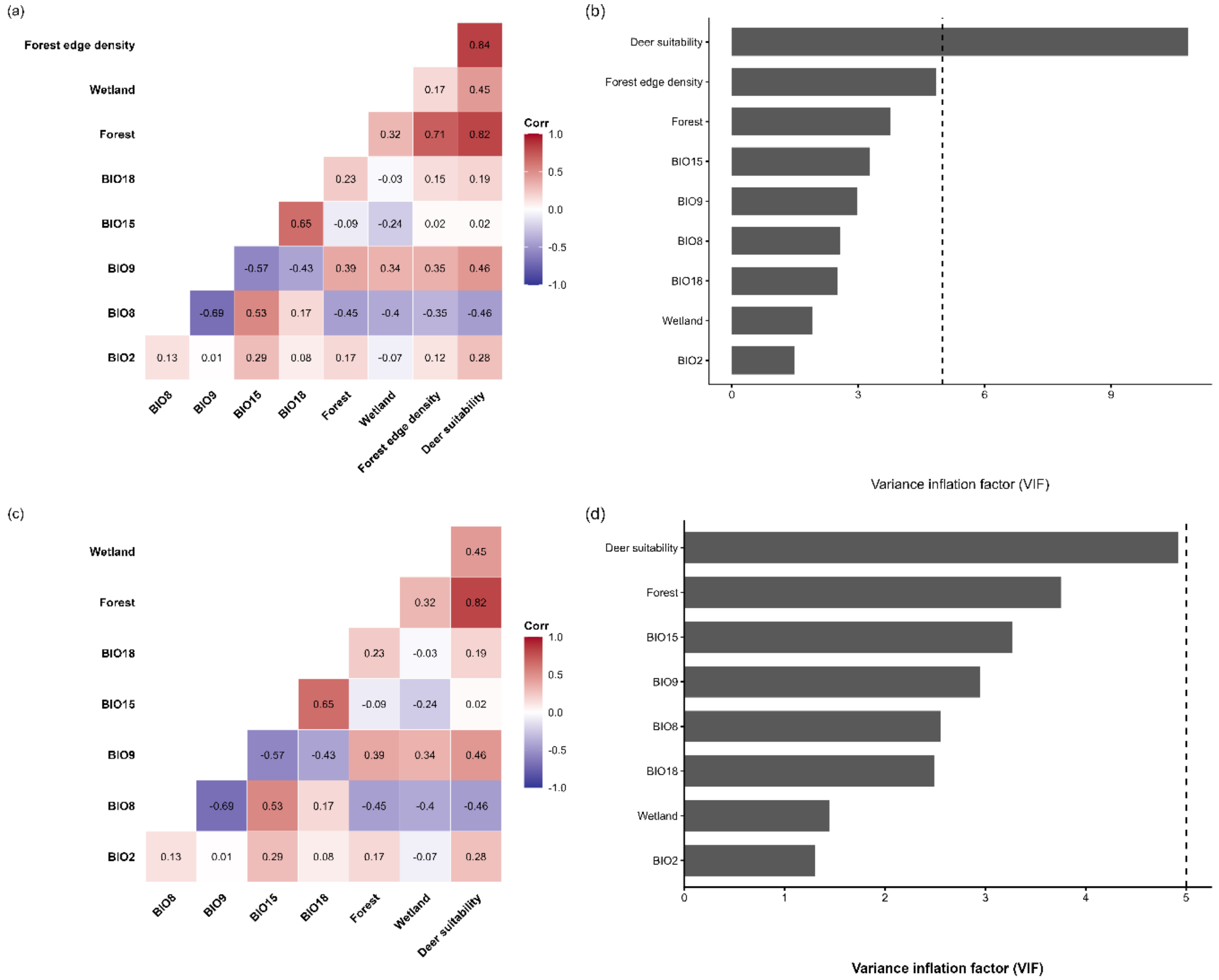


**Figure S1.** Multicollinearity assessment of county-level ecological predictors. (a) Pairwise Pearson correlation coefficients among the nine initial candidate predictors. (b) Variance inflation factors (VIFs) for the nine candidate predictors. (c) Pairwise Pearson correlation coefficients among the eight retained predictors after excluding forest-edge density. (d) VIFs for the eight retained predictors. The dashed horizontal line indicates a VIF threshold of 5.

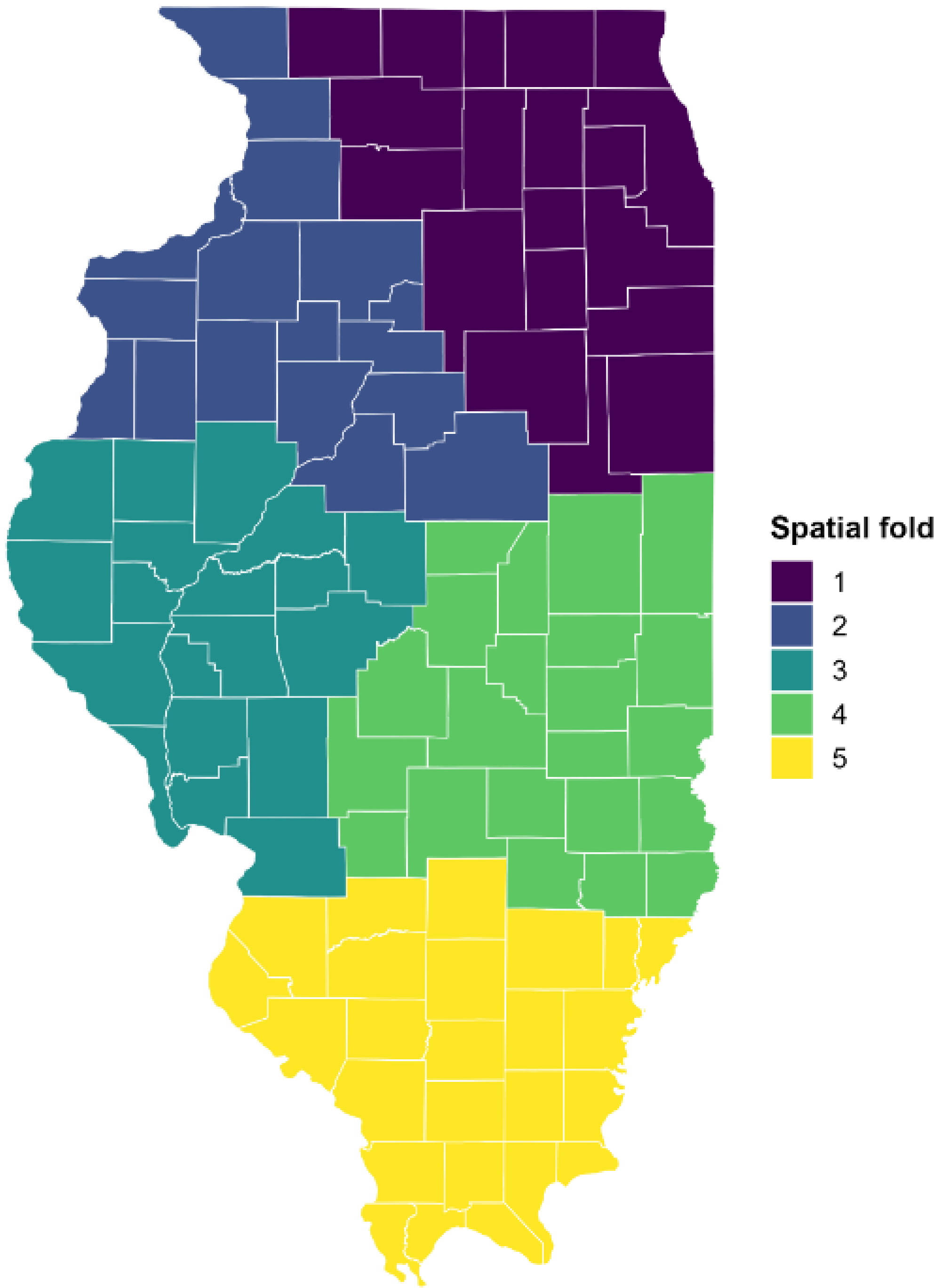


**Figure S2.** Geographic distribution of five spatial cross-validation folds across 102 Illinois counties, generated using k-means clustering of projected county coordinates (EPSG:5070).

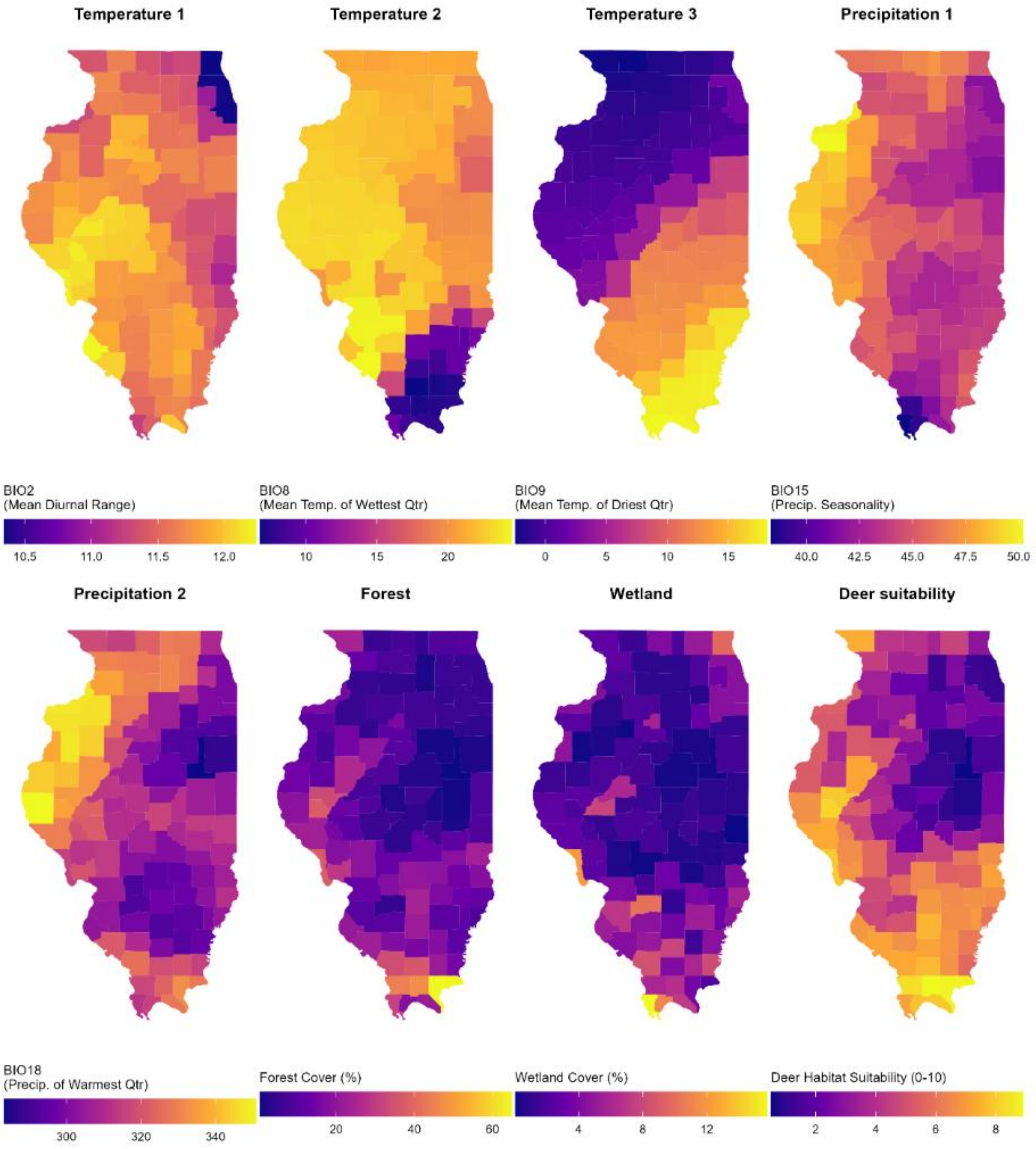


**Figure S3.** County-level geographic distribution of eight ecological predictors across Illinois, including five bioclimatic variables (BIO2, BIO8, BIO9, BIO15, and BIO18), forest cover, wetland cover, and white-tailed deer habitat suitability.

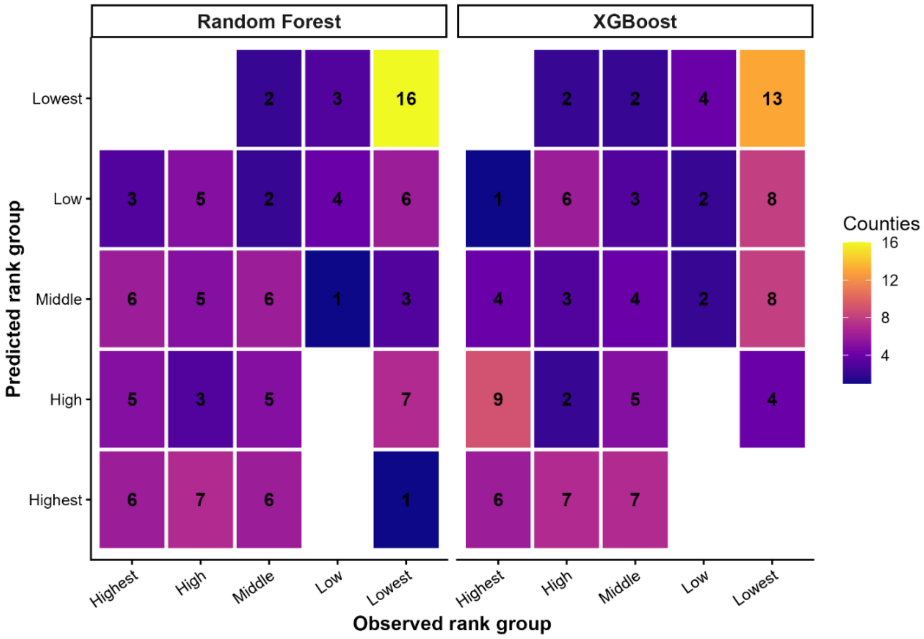


**Figure S4.** Agreement between observed and spatially cross-validated predicted lone star tick-proportion rank groups across 102 Illinois counties. Heatmaps show county counts for the random forest and XGBoost across five descending-rank groups. Tied observations received average ranks, resulting in unequal group sizes, including 33 counties with zero observed lone star ticks in the lowest group.

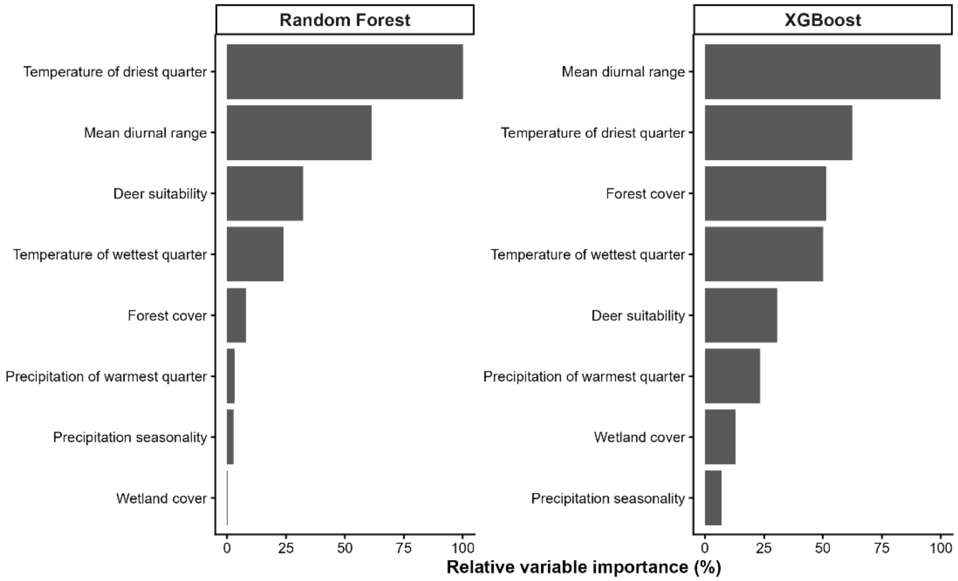


**Figure S5.** Relative importance of ecological predictors in random forest and XGBoost models of lone star tick relative abundance across Illinois. Predictor importance was estimated using permutation importance for random forest and gain for XGBoost. Values were normalized separately within each model, with the highest-ranked predictor assigned an importance of 100%.

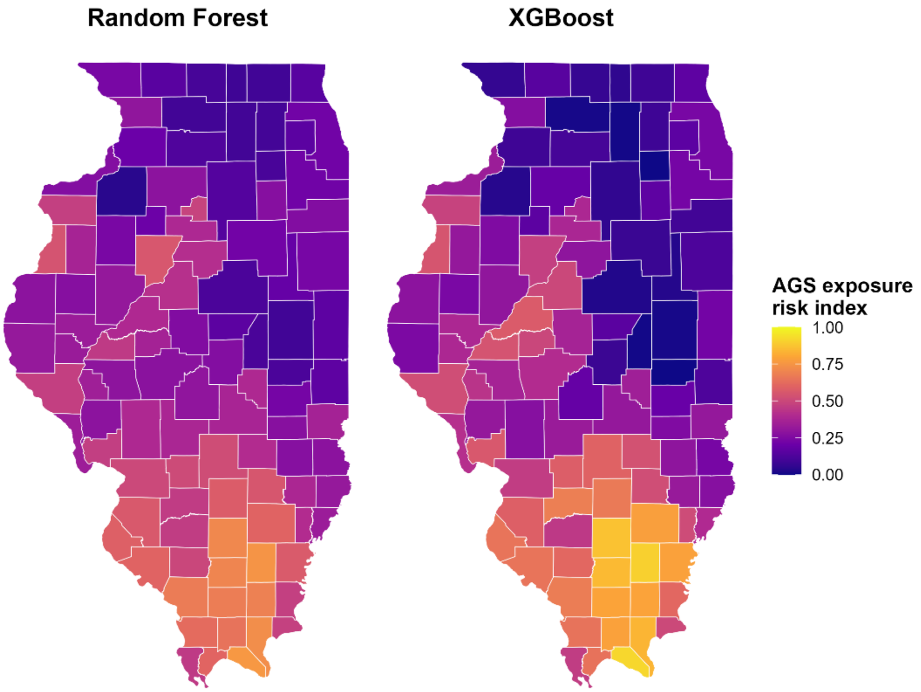


**Figure S6.** Geographic distribution of WUI-adjusted alpha-gal syndrome exposure-risk indices. Random forest and XGBoost indices incorporate predicted lone star tick proportions (70%), scaled ehrlichiosis incidence (20%), and wildland-urban interface population (10%).

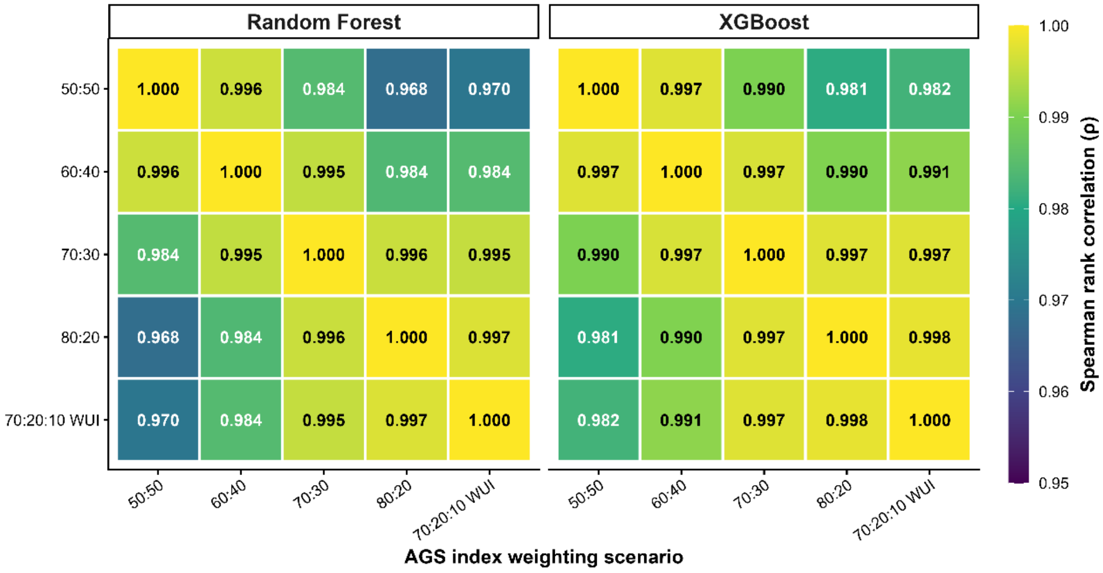


**Figure S7.** Agreement among alternative alpha-gal syndrome exposure-risk index formulations. Pairwise Spearman correlations compare four ecological-to-ehrlichiosis weighting scenarios and the WUI-adjusted formulation across 102 Illinois counties, separately for random forest and XGBoost.

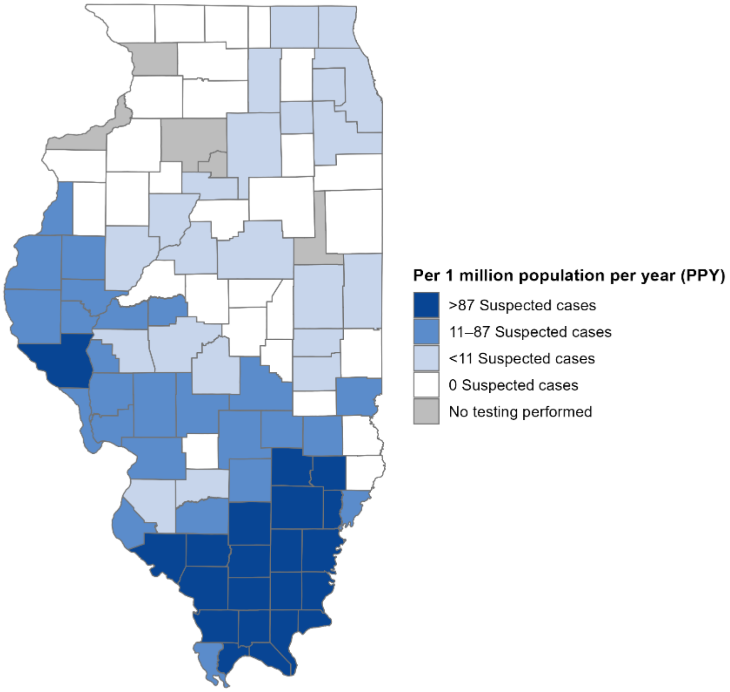


**Figure S8.** Geographic distribution of CDC suspected-AGS categories across Illinois. County-level categories represent reported suspected-AGS cases per million population annually during 2017-2022.

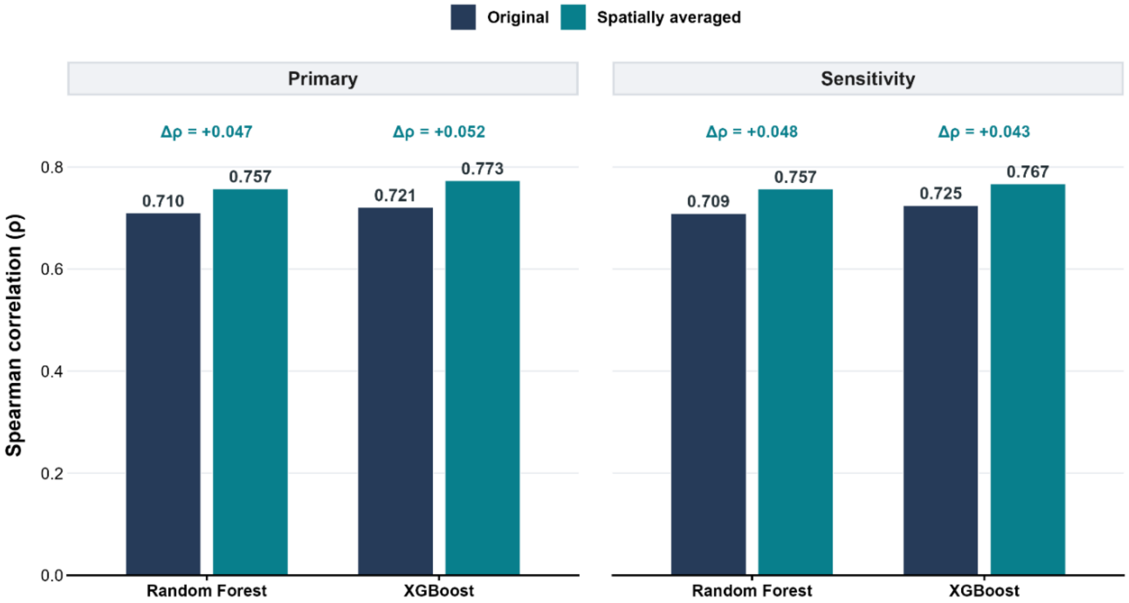


**Figure S9.** Effect of spatial averaging on agreement with CDC suspected-AGS categories. Spearman correlations compare the original and spatially averaged primary and WUI-adjusted indices for the random forest and XGBoost models.
